# Multicenter evaluation of the Panbio™ COVID-19 Rapid Antigen-Detection Test for the diagnosis of SARS-CoV-2 infection

**DOI:** 10.1101/2020.11.18.20230375

**Authors:** Paloma Merino-Amador, Jesús Guinea, Irene Muñoz-Gallego, Patricia González-Donapetry, Juan-Carlos Galán, Nerea Antona, Gustavo Cilla, Silvia Hernáez-Crespo, José-Luis Díaz-de Tuesta, Ana Gual-de Torrella, Fernando González-Romo, Pilar Escribano, Miguel Ángel Sánchez-Castellano, Mercedes Sota-Busselo, Alberto Delgado-Iribarren, Julio García, Rafael Cantón, Patricia Muñoz, M Dolores Folgueira, Manuel Cuenca-Estrella, Jesús Oteo-Iglesias, Spanish Panbio™ COVID-19 validation group ‡

**Affiliations:** Microbiology Department, Hospital Universitario Clínico San Carlos, Madrid, Spain; Microbiology Department, Hospital Universitario Gregorio Marañón, Madrid, Spain; Clinical Microbiology Department, Hospital Universitario Doce de Octubre, Madrid, Spain; Microbiology Department, Hospital Universitario La Paz, Madrid, Spain; Microbiology Department, Hospital Universitario Ramón y Cajal e Instituto Ramón y Cajal de Investigación Sanitaria (IRYCIS), Madrid, Spain; CIBER de Investigación en Salud Pública (CIBERESP). Madrid. Spain; Osakidetza Basque Health Service, Hospital Universitario Cruces, Microbiology Department, Barakaldo, Spain; Health Research Institute, Biodonostia, San Sebastián, Spain; Osakidetza Basque Health Service, Hospital Universitario Donostia, Microbiology Department, San Sebastián, Spain; Osakidetza Basque Health Service, Hospital Universitario Araba, Microbiology Department, Vitoria, Spain; Osakidetza Basque Health Service, Hospital Universitario Basurto, Microbiology Department, Bilbao, Spain; Osakidetza Basque Health Service, Hospital Universitario Galdakao-Usansolo, Microbiology Department, Galdakao, Spain; Osakidetza Basque Health Service, Hospital Universitario Donostia, Clinical Management Unit of Gipuzkoa Laboratories, San Sebastián, Spain; Spanish Network for Research in Infectious Diseases (REIPI); Instituto de Investigación Hospital 12 de Octubre, imas12, Madrid, Spain; Department of Medicine, Universidad Complutense School of Medicine, Madrid, Spain; Instituto de Salud Carlos III, Majadahonda, Madrid, Spain; National Centre for Microbiology, Instituto de Salud Carlos III, Majadahonda, Madrid, Spain

**Keywords:** COVID-19, rapid antigen-detection test, SARS-CoV-2

## Abstract

The standard RT-PCR assay for COVID-19 is laborious and time-consuming, limiting the availability of testing. Rapid antigen-detection tests are faster and less expensive; however, the reliability of these tests must be validated before they can be used widely. The objective of this study was to determine the reliability of the Panbio^TM^ COVID-19 Ag Rapid Test Device (PanbioRT) (Abbott) for SARS-CoV-2 in nasopharyngeal swab specimens. This was a prospective multicenter study in ten Spanish university hospitals of patients from hospital units with clinical symptoms or epidemiological criteria for COVID-19. Patients whose onset of symptoms or exposure was more than 7 days earlier were excluded. Two nasopharyngeal exudate samples were taken to perform the PanbioRT and a diagnostic RT-PCR test. Among the 958 patients studied, 359 (37.5%) were positive by RT-PCR and 325 (33.9%) were also positive by the PanbioRT. Agreement was 95.7% (kappa score: 0.90). All 34 false-negative PanbioRT results were in symptomatic patients, with 23.5% of them at 6–7 days since the onset of symptoms and 58.8% presenting *C*_*T*_ >30 values for RT-PCR, indicating a low viral load. Overall sensitivity and specificity for the PanbioRT were 90.5% and 98.8%, respectively. The PanbioRT provides good clinical performance as a point-of-care test, with even more reliable results for patients with a shorter clinical course of the disease or a higher viral load. While this study has had a direct impact on the national diagnostic strategy for COVID-19 in Spain, the results must be interpreted based on the local epidemiological context.

## Introduction

The novel coronavirus SARS-CoV-2, which causes severe acute respiratory syndrome, emerged in Wuhan (China) in December 2019. The virus has spread rapidly, causing a global pandemic that was defined by The World Health Organization (WHO) as Coronavirus disease-19 (COVID-19) (1). Reliable and early diagnosis of the responsible agent, not only in symptomatic patients but also in close contacts of confirmed cases, is critical to controlling the spread of the disease (2,3). Clinical diagnosis of COVID-19 has relied on nucleic acid amplification tests, such as real-time reverse transcription polymerase chain reaction (RT-PCR) assays to detect SARS-CoV-2 (3,4). RT-PCR, the current standard for COVID-19 diagnosis, is also the standard test in clinical microbiology laboratories for the diagnosis of many infections because the test is sensitive and pathogen-specific. However, the method is laborious, requiring inactivation of the virus and RNA extraction before the RT-PCR assay. The test (under optimal conditions) takes approximately 4-5 h, and the response time is typically 12 to 24 h. This test also requires a continuous supply of PCR kits as well as swabs and transport media for samples, inactivation solutions, extraction reagents, and disposable plasticware. The WHO recommended at least 200 PCR tests per 100,000 population to ensure adequate control of the pandemic; however, in Spain more than 1,500 PCR tests per 100,000 population were performed in September and October 2020 (5). This high level of testing will be difficult to maintain in winter because of testing for other respiratory infections, such as influenza. Therefore, we risk saturating the PCR-testing capacity of microbiology laboratories, but also of sampling capacities in primary care centers.

There are ongoing efforts to develop fast, reliable, and inexpensive diagnostic tests specific for detection of SARS-CoV-2 antigens. Rapid antigen-detection tests (RADT) for both laboratory and near-patient use detect SARS-CoV-2 proteins produced by replicating viruses in respiratory secretions (6). The Panbio^TM^ COVID-19 Ag Rapid Test Device (PanbioRT) (Abbott Diagnostic GmbH, Jena, German) is a rapid in vitro test for the qualitative detection of the viral nucleocapsid protein in nasopharyngeal swab specimens from individuals who meet clinical and/or epidemiological criteria of COVID-19 infection. This is a novel, lateral-flow-format test that uses immunochromatography with colloidal gold in a point-of-care format, with results in 15-20 min. The objective of this study is to determine the reliability of the PanbioRT with CE marking.

## Materials and Methods

This was a prospective multicenter diagnostic evaluation study across ten independent university hospitals in two Spanish autonomous communities (Madrid and Basque Country) using consecutive enrollment. The hospitals participating in the study from Madrid were Hospital Clínico Universitario San Carlos, Hospital Universitario Ramón y Cajal, Hospital Universitario La Paz, Hospital Universitario Doce de Octubre, and Hospital Universitario Gregorio Marañón. Hospitals from Basque Country were Hospital Universitario Araba, Hospital Universitario Cruces, Hospital Universitario Basurto, Hospital Universitario Donostia, and Hospital Universitario Galdakao-Usansolo. Patients with clinical symptoms or epidemiological criteria (asymptomatic close contacts) for COVID-19 from hospital emergency rooms or other hospital units who were to receive a diagnostic RT-PCR test were included in the study. All participants were reported as part of the study and verbal informed consent was obtained prior to their inclusion. The result of the PanbioRT did not influence the clinical management of the patients, which was decided based on the RT-PCR result. Patient data were coded and no samples were stored after the PanbioRT was performed. Patients whose onset of symptoms or exposure was more than 7 days earlier were excluded.

Two nasopharyngeal exudate samples were taken per patient: one was used immediately by trained personnel to perform the PanbioRT according to the manufacturer’s instructions, and the second was used for a molecular diagnostic (RT-PCR) by each hospital according to its standard procedures for COVID-19 diagnosis.

The symptoms, number of days since the onset of symptoms or exposure, threshold cycle (*C*_*T*_) values for PCR, and demographic data were collected for all participants. Specificity and sensitivity with 95% confidence intervals (CI) were calculated using the RT-PCR results as the standard. Sensitivity was calculated for all patients and for specific groups of patients according to the time of onset of symptoms or exposure, RT-PCR *C*_*T*_ values, symptoms, and age. The level of agreement between the tests was evaluated using Cohen’s kappa score (7).

## Results

Between September and October 2020, 958 individuals who had at least one symptom compatible with COVID-19 (n=830) or had been in close contact with a diagnosed COVID-19 patient (n=128) were included in this study. There were between 8 and 245 individuals from each participating hospital with a mean age of 42.4 years (range, 1-100); 61.3% were women (Table 1) and 58 cases were in pediatric patients (≤14 years old).

**Table 1.** Study cohort included in the validation study of Panbio^TM^ COVID-19 Ag Rapid Test Device.

|  |  |
| --- | --- |
| Total N (valid PCR results) | **958** |
| Positive PCR [% (n)] | **37.5% (359)** |
| Age [mean (min–max)] | **42.4 (1–100)** |
| Gender [% F, (n/N)] | **61.3% (587/958)** |
| Symptoms present [% Yes, (n/N)] | **86.6% (830/958)** |
| Days from symptom onset or from exposure [mean (N)] | **2.8 (958)** |
| Days ≤5 [n/N (%)] | **854/958 (89.1%)** |
| Days 6–7 [(n/N (%)] | **104/958 (10.9%)** |
| PCR $C_T$ (n) | **295** |
| $C_T \geq 30$ [n/N, (%)] | **41 (13.9%)** |
| $C_T < 30$ [n/N, (%)] | **254 (86.1%)** |

Among these 958 patients, 359 (37.5%) were positive by RT-PCR and 325 (33.9%) were also positive by the PanbioRT (Table 2). A total of 599 (62.5%) were negative by PCR and 592 (61.8%) were also negative by the PanbioRT (Table 2). The agreement between both methods was 95.7% (kappa score: 0.90; CI 95%: 0.88–0.93). In 41 patients the results differed between the two tests and most of the differences were in individuals who tested positive with the RT-PCR test but negative with the PanbioRT (n=34; 3.5%) (Table 2, Table 3). All 34 of these cases were in symptomatic patients, with 8 (23.5%) of them at 6–7 days since the onset symptoms, and 20 (58.8%) had C_T_>30 values for RT-PCR.

**Table 2.** Summary of the results of the Panbio^TM^ COVID-19 Ag Rapid Test Device compared to RT-PCR.

|  | **Panbio™ COVID-19 Ag Rapid Test Device** |  |  |
| --- | --- | --- | --- |
| **RT-PCR** | Positive | Negative | TOTAL |
| Positive | 325 | 34 | 359 |
| Negative | 7 | 592 | 599 |
| TOTAL | 332 | 626 | 958 |

**Table 3.** Cases with discordant results between the RT-PCR method and the PanbioRT.

| **Sex** | **Compatible symptoms** | **Days since symptoms onset** | **Panbio result** | **RT-PCR result** | **C<sub>T</sub> RT-PCR** |
| --- | --- | --- | --- | --- | --- |
| Male | Yes | 1 | Negative | Positive | 27 |
| Female | Yes | 1 | Negative | Positive | 34 |
| Male | Yes | 5 | Negative | Positive | 25 |
| Male | Yes | 6 | Negative | Positive | 29 |
| Female | Yes | 6 | Negative | Positive | 31 |
| Male | Yes | 5 | Negative | Positive | 26 |
| Female | Yes | 1 | Negative | Positive | 32 |
| Female | Yes | 2 | Negative | Positive | 37 |
| Female | Yes | 1 | Negative | Positive | 28 |
| Male | Yes | 3 | Negative | Positive | 32 |
| Male | Yes | 1 | Negative | Positive | 32 |
| Female | Yes | 7 | Negative | Positive | 30 |
| Male | Yes | 6 | Negative | Positive | 31 |
| Female | Yes | 2 | Negative | Positive | 26 |
| Female | Yes | 2 | Negative | Positive | 36 |
| Female | Yes | 1 | Negative | Positive | 30 |
| Female | Yes | 3 | Negative | Positive | 37 |
| Female | Yes | 5 | Negative | Positive | 35 |
| Female | Yes | 3 | Negative | Positive | 29 |
| Female | Yes | 3 | Negative | Positive | 26 |
| Female | Yes | 7 | Negative | Positive | 25 |
| Female | Yes | 2 | Negative | Positive | 26 |
| Female | Yes | 4 | Negative | Positive | 31 |
| Female | Yes | 3 | Negative | Positive | 34 |
| Male | Yes | 1 | Negative | Positive | 34 |
| Female | Yes | 7 | Negative | Positive | 38 |
| Male | Yes | 3 | Negative | Positive | 34 |
| Female | Yes | 4 | Negative | Positive | 34 |
| Female | Yes | 7 | Negative | Positive | 38 |
| Male | Yes | 3 | Negative | Positive | 34 |
| Male | Yes | 1 | Negative | Positive | 32 |
| Female | Yes | 3 | Negative | Positive | 37 |
| Female | Yes | 5 | Negative | Positive | 28 |
| Male | Yes | 7 | Negative | Positive | 30 |
| Female | Yes | 6 | Positive | Negative |  |
| Male | Yes | 7 | Positive | Negative |  |
| Female | Yes | 1 | Positive | Negative |  |
| Female | Yes | 1 | Positive | Negative |  |
| Male | Yes | 1 | Positive | Negative |  |
| Female | Yes | 3 | Positive | Negative |  |
| Female | Yes | 5 | Positive | Negative |  |

Based on these data, the overall sensitivity and specificity of the PanbioRT were 90.5% (CI 95%: 87.5–93.6) and 98.8% (CI 95%: 98–99.7), respectively (Table 3). Sensitivity was slightly higher in patients with ≤5 days of the clinical course of the disease (91.8%; CI 95%: 88.8–94.8) or in those who had a C_T_ <30 for the RT-PCR test (94.5%; CI 95%: 91.7–97.3) (Table 4).

**Table 4.** Estimation of clinical performance of the Panbio^TM^ COVID-19 Ag Rapid Test Device compared to RT-PCR.

|  |  |
| --- | --- |
| Relative Sensitivity (CI 95%) | 90.5% (87.5–93.6) |
| Sensitivity days $\leq 5$ (CI 95%) | 91.8% (88.8–94.8) |
| Sensitivity $C_T < 30$ (CI 95%) | 94.5% (91.7–97.3) |
| Relative Specificity (CI 95%) | 98.8% (98–99.7) |
| Agreement (kappa index; CI 95%) | 95.7% (0.9; 0.88–0.93) |
| Positive predictive value (CI 95%) | 97.8% (96.3–99.4) |
| Negative predictive value (CI 95%) | 94.6% (92.8–96.3) |

Among the 128 asymptomatic participants who had close contact with a COVID-19 patient, there was full concordance in the 31 (24.2%) who were positive by RT-PCR and in the 97 that were negative. Six (10.3%) of the 58 pediatric patients included in the study were positive by RT-PCR and also by the PanbioRT.

The negative predictive value (NPV) and positive predictive value (PPV) in the study cohort, with a high prevalence (37.5%), were 94.6% and 97.8%, respectively (Table 4). PPV and NPV were also calculated for lower prevalence of 5% and 10%; the results obtained were 79.8% and 89.3%, respectively, for PPV; and 99.5 and 98.9%, respectively, for NPV.

## Discussion

As the COVID-19 pandemic continues unabated, the gap between the number of tests that are needed and the testing capacity of laboratories or in primary care settings increases (3). RADT tests are simple to perform and interpret without the need for equipment at the point-of-care, they are inexpensive, and they provide quick results. Early rapid antigen-detection test for COVID-19 diagnosis had poor reliability that precluded their general use (8-11); however, the new generation of RADT appear to have substantially improved reliability (12-15). Although these rapid tests show promise for use as part of a larger strategy for COVID-19 diagnosis and control (6), there are insufficient validation studies to support their use in varied patient environments. In this study the PanbioRT gave very good clinical performance values, with 90.5% sensitivity and 98.8% specificity; moreover, sensitivity was even improved in patients with five or fewer days of clinical progression of the disease. The strengths of this study include the large study size, the high percentage of positive cases, the inclusion of multiple centers, the prospective nature of the study, and the inclusion of point-of-care patients. This study has had an immediate clinical impact, having been used to modify the Spanish Strategy for Early Detection, Surveillance, and Control, of COVID-19 (Update September 25, 2020) (16).

In two recent pre-published Spanish studies with 412 patients (54 positive by RT-PCR) (13) and 255 patients (60 positive by RT-PCR) (12), the overall sensitivities were 79.3% and 76.3%, respectively; however, in the second study sensitivity was 86.5% in symptomatic patients with seven or fewer days since the onset of symptoms (12). WHO guidelines require that SARS-CoV-2 RADT demonstrate ≥80% sensitivity and ≥97% specificity compared to the RT-PCR reference assay (6). Thus, our data support the clinical use of the PanbioRT instead of the RT-PCR test in patients with symptoms of COVID-19 with a short clinical course (≤5–7 days) of the disease. Although the results obtained in asymptomatic patients and children under 14 years of age were good, the number of cases included for these subpopulations was small (128 and 58, respectively), making it inadvisable to conclude general results about that. In a pre-published study with frozen samples (17), sensitivity was significantly higher among samples collected in the setting of case identification (92.6%) and contact tracing (94.2%) than in asymptomatic screening (79.5%). This is consistent with the advice from the WHO against using RADT for screening asymptomatic individuals in populations with low COVID-19 prevalence (6) due to the potential increase of higher incidence of false positives. In this study, we estimated that the PPV decreased to 79.8% when the prevalence was 5%.

The performance of an RADT may depend on the epidemiological situation of the population being tested; therefore, how the test is used and how the results are interpreted will depend on local epidemiological factors (6). In populations with a high prevalence and a high frequency of symptomatic patients, a positive rapid test would be considered confirmatory for infection. However, a negative result would lead to further testing for respiratory pathogens, including an RT-PCR test for COVID-19 if the symptoms were consistent with this disease. In populations with a low prevalence of COVID-19 and more asymptomatic patients, a negative test would be accepted, but a positive test, which is more likely to be false, could require a confirmatory RT-PCR test.

The use of RADT as a diagnostic tool can greatly reduce the testing burden on microbiology laboratories. However, in the primary care setting, which has also reached saturation in the testing and diagnosis of COVID-19, changes would be required to allow them to perform the rapid test on-site. The ability to perform this test in patient care centers would simplify the process of testing, provide rapid results to the doctor and the patient, thus improving the decision-making process and reducing pressure on the health care providers.

In conclusion, this study showed that the PanbioRT provides very good clinical performance as a point-of-care test, with even better results for patients with a shorter clinical course of the disease or higher viral load. While this study has had a direct impact on the national diagnostic strategy for COVID-19 in Spain, the results must be interpreted based on the local epidemiological context. The ease and speed of RADT with god clinical performance could help to prevent an overload on health care services as laboratories will have to cope with an increase in respiratory infections during winter.

## Data Availability

All data mentioned in the manuscript are available upon request to the corresponding author

## Acknowledgments

We thank Abbott Diagnostics for providing Panbio™ COVID-19 AG Rapid Test Device kit and all the professionals involved in the diagnosis of COVID-19 in the participating hospitals.

Members of the Spanish Panbio™ COVID-19 validation group are:

Sara Medrano and Alba Pérez (H. Universitario Clínico San Carlos), Oscar Martínez-Expósito (H. Universitario Cruces), Izaskun Alejo-Cancho (H. Universitario Galdakao), M. Carmen Martín-Higuera and Marta Rolo (H. Universitario Doce de Octubre), M^a^ Jesús Estévez and Tania Bravo (H. Universitario Ramón y Cajal), Diego Vicente and Mila Montes (H. Universitario Galdakao-Usansolo).

